# Virtual Neuronavigation for Parcel-guided TMS

**DOI:** 10.1101/2025.10.14.25337524

**Authors:** Dennis Q Truong, Abhishek Datta, Jeffrey Moreno, Yishai Z Valter, Rashel Mejia, Kamran Nazim, Yifan Gao, Yu Huang, Joshua A Berman, Daniel C Javitt

**Affiliations:** Research and Development, Soterix Medical, Inc, Woodbridge, NJ; City College of New York, New York, NY; Columbia University, New York, NY

**Keywords:** rTMS for depression, treatment resistant depression, HCP atlas, neuronavigation, coil positioning, individualized positioning

## Abstract

**Background:** rTMS for depression is partially effective which is likely in part due to the treatment coil positioning being based on scalp-based approaches. Such an approach results in not accounting for *individual* anatomy and therefore delivering electrical currents to non-optimal brain targets. In a pilot study, we demonstrated that rTMS targeted to parcel 46 (p46) of the HCP atlas led to 100% response in patients who were resistant to the standard TMS. Implementing parcel-guided targeting requires neuronavigation. However, neuronavigation use in clinical practice is limited. Besides added cost, need for individual MRI, leveraging new targeting advances requires expertise in 3D brain reconstruction and processing.

**Methods:** We sought to develop a virtual neuronavigation system in which MRI images are processed on the backend to determine p46 and a corresponding individualized 3D printed scalp headgear is developed. The headgear includes markings for the location corresponding to p46 and other anatomical guides. Headgear development involved an iterative sequence of 3D CAD modeling, printing, and testing to meet the gold standard neuronavigation-guided location. Individual headgear accuracy is then verified by positioning on the intended subject and comparing alignment with the location on the scalp and brain intersecting the neuronavigation beam targeting p46. Reproducibility testing was also performed.

**Results:** Across 16 test subjects on which the final version of the headgear could be tested, individualized headgear was found to be 8.31± 4.93 mm from the neuronavigation-determined scalp location. The corresponding distances on the brain was 7.94± 4.99 mm. Within-subject reproducibility (against centroid formed by 3 measurements) was 1.21 ± 0.84 mm across n= 8 subjects.

**Conclusion:** Individualized headgear targeting presents a viable option for practices to leverage state-of-the-art targeting advances without investing in neuronavigation hardware.

## Introduction

Repetitive transcranial magnetic stimulation (rTMS) is an FDA-approved treatment for conditions including treatment-resistant major depression (TRD), anxiety, obsessive-compulsive disorder (OCD), and smoking cessation (O’Reardon 2007). In the context of depression, the treatment involves positioning an electromagnetic coil above the scalp to induce therapeutic electrical currents in the left dorsolateral prefrontal cortex (lDLPFC). The original FDA-approved protocol, however, utilizes a scalp-based “5-cm rule” for coil placement, which inherently overlooks individual anatomical variability. Given that brain imaging techniques were relatively underdeveloped at the time of FDA approval in 2008, a scalp-based approach was considered a practical solution.

Currently, rTMS exhibits a response rate of approximately ∼45% (Connolly 2012, Carpenter 2012, Blumberger 2018), a limitation that may stem partly from the reliance on nomothetic targeting methods that do not account for interindividual differences in brain and scalp anatomy. Notably, the “5-cm rule” has been found to deviate from the precise location of the lDLPFC by several millimeters (George 2010, Herwig 2001). While alternative scalp-based strategies—such as the 5.5-cm rule (George 1995) and the EEG F3 method (Jasper 1958)—have been proposed to enhance targeting accuracy, they still fall short of incorporating brain-based personalization. Consequently, there is a growing need for individualized approaches to optimize treatment outcomes.

In recent years, evidence has increasingly demonstrated that TMS targeting anhedonic symptoms in TRD is most effective when aimed at the portion of the lDLPFC that is maximally anti-correlated with the subgenual anterior cingulate cortex (sgACC), as determined by resting-state functional connectivity MRI (rs-fMRI). However, identifying this optimal target requires access to fMRI—a resource often limited to specialized research institutions—and the use of complex processing pipelines that demand significant technical expertise. Moreover, patient-specific targeting typically employs neuronavigation systems that co-register the subject’s MRI with the TMS coil in a virtual environment, using magnetic or optical tracking to provide real-time positional feedback. The high cost of neuronavigation hardware and the requisite technical skills have hindered widespread clinical adoption.

A more recent “surface-based” brain reconstruction approach, developed as part of the Human Connectome Project (HCP) (Glasser, 2016), requires only structural scans rather than functional imaging. This method leverages the greater consistency of the two-dimensional cortical surface across individuals compared to the three-dimensional brain volume, thereby reducing the extent of smoothing required when aligning analogous regions across subjects. In this framework, the cortical surface is independently rendered from underlying structures into a network of vertices, or “grayordinates,” which are then categorized into distinct regions and parcels according to the first-generation HCP Multimodal Parcellation (HCP MMP1.0) atlas (Van Essen, 2013). Notably, while the traditional Brodmann map divides the brain into 43 regions, the HCP MMP1.0 atlas partitions it into 180 distinct parcels. Moreover, the lDLPFC is segmented into 13 parcels, with Brodmann areas 9 and 46 further subdivided into four distinct parcels. Functional connectivity between parcel 46 and the subgenual anterior cingulate cortex (sgACC), along with other brain regions, has been correlated with clinical improvement following effective electroconvulsive therapy (ECT), underscoring the therapeutic relevance of this target in depression treatment (Moreno-Ortega, 2019).

A recent study compared outcomes in two cohorts—10 TMS-resistant patients who received rTMS targeted at parcel 46 and 22 TMS-naïve patients treated with standard TMS (Moreno-Ortega, 2020). All TMS-resistant individuals exhibited significant clinical response, with 50% achieving remission, whereas only 45% of the TMS-naïve group responded to the standard protocol. Parcel-guided rTMS thus presents the possibility of utilizing surface-based reconstruction to precisely locate an optimal cortical target without relying on fMRI, thereby enhancing the accessibility of personalized rTMS.

For parcel-guided rTMS to be widely implemented amongst TMS providers, however, it is essential to develop targeting methods that do not depend on neuronavigation hardware. The challenge is compounded by the rising interest in remote rTMS administration—an interest that has surged since the COVID-19 pandemic (Caulfield, 2020) and is further supported by advances in miniaturized TMS coils (Lee, 2019). In response to these challenges, we sought to develop a pipeline for enabling parcel-guided rTMS without neuronavigation hardware. Our approach comprises precise localization of parcel 46, automated segmentation of the subject’s structural scan, and the design of customized headgear that guides clinicians to the optimal scalp location for TMS coil placement.

## Methods

### Participants

Individual MRI was obtained as part of ongoing trials (ClinicalTrials.gov IDs: NCT05598931 and NCT04956081). Male and female subjects (n = 35, 10 female) ages 21-66 years were recruited. All subjects gave written informed consent for study participation and ethical approval was obtained from the WCG IRB (approval #20213876). Due to constraints with participant availability over the course of headgear development, accuracy of the final headgear version could only be tested on a limited subset of subjects (n=16, 4 female).

### MRI acquisition

Subjects underwent MRI on a Siemens 3T Prisma scanner equipped with 32 channel head coil. Structural MRI scans consistent with the HCP were acquired (Glasser 2016). These sequences consisted of two T1-weighted MPR vNav scans at 0.8 mm isotropic, matrix = 320x300, slices = 208, TR = 2500 ms, TE = 3.02 ms, flip angle = 8° and two T2-weighted SPC vNav scans at 0.8 mm isotropic, matrix = 320x300, slices = 208, TR = 3200 ms, TE = 564 ms, flip angle = 120°. See **Figure 1A** for the overall manufacturing process of the individualized headgear starting with structural scans.

**Fig. 1.**
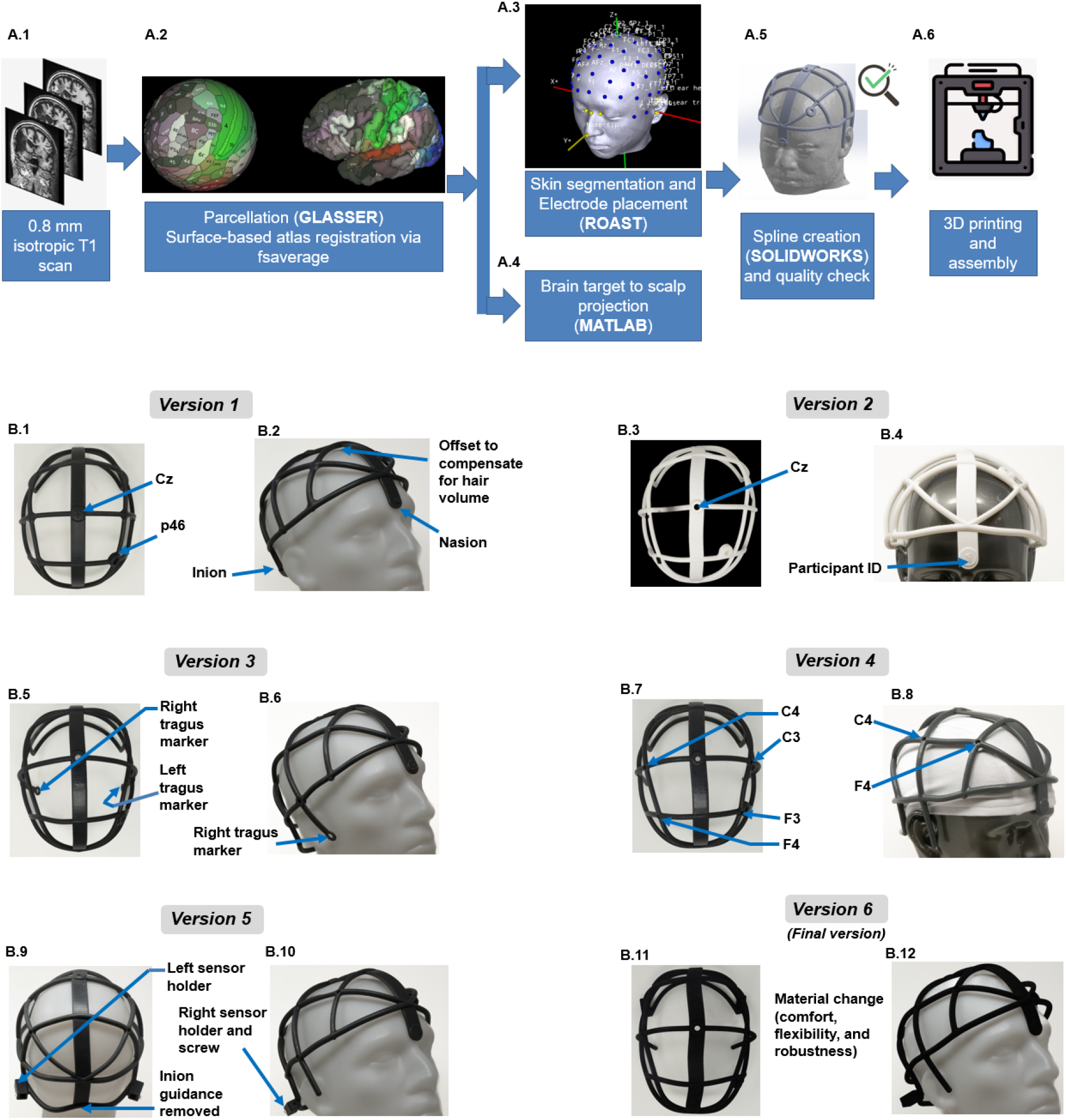
Development of the individualized head gear and manufacturing process of the final version. Top row. An unique 6-stage development process was developed to generate a personalized headgear that allows marking the scalp coordinate corresponding to p46. **Bottom row**. A typical iterative design workflow involving ideation, 3D printing, and testing was followed with the goal to match neuronavigation-guided prediction. See Methods for details. The 6 major design iterations with main functional elements are shown here.

### Determination of brain target

The cortical surface of each subject was reconstructed using the function ‘recon-all’ (Freesurfer v6.0.0) with the structural MRI sequences as input. Individual parcellation was obtained by registering the HCPMMP1 atlas labels (Glasser 2016) to each subject’s native space surface using the ‘fsaverage’ surface as an intermediary (https://github.com/tannerjared/HCP-MMP1). The parcel was subsequently mapped from the surface to a volumetric segmentation mask for loading into the navigation software as well the headgear production pipeline (**Figure 1A.4**).

### Determination of virtual scalp target

In order to target a brain location without neuronavigation, we developed a stereotactic headgear. This requires projection of the brain target to the scalp surface. Several methods were evaluated for virtually projecting a brain target to the scalp: nearest, normal, and radial. Prior to manufacturing the headgear, a subset of 10 initial subjects were evaluated with each method. The nearest method calculated the nearest voxel of the skin surface to the target. The normal method starts with the nearest scalp location then iteratively scans the surrounding surface normals of the scalp mesh to find the closest intersecting surface normal to the target. The radial method projects a line from the MNI origin through the brain target to the skin surface. The worldspace coordinate of each method was calculated and compared to the measured scalp location when aiming a coil at the corresponding brain target.

### Individual headgear development

An *iterative* design workflow under the medical device development framework was followed. The process was initiated by setting an extensive list of design inputs ranging from functional utility, manufacturability, comfort, usability, etc. It was clear that a whole-head personalized headgear with mesh-like support elements would be needed for this objective. We decided to leverage an open-source transcranial electrical stimulation modeling software (ROAST) developed by members of our team (Huang 2017) to obtain 10-20 electrode coordinate locations guided by participant MRI. Specifically, skin tissue was first segmented and electrode coordinate locations identified to ensure locations were on top of the tissue mask (i.e. not submerged and ∼1 mm above the skin surface). In parallel, the projection from the p46 brain location onto the subject’s scalp was also available as input to the headgear creation process (**Figure 1A.4)**. With respect to headgear design, sketches realizing initial design inputs were first drafted followed by an iterative sequence of 3D CAD modeling, 3D printing, assembly, and subsequent testing on participants (Valter 2021a; Valter 2021b). With every iteration, the goal was to reduce the distance to the gold standard (i.e. the individual MRI-neuronavigation guided location).

The STL file for the scalp mask, 10-20 coordinate locations, and the identified scalp location corresponding to p46 were fed to the CAD software. Using an in-house script, spline segments were automatically created from each of the 10-20 coordinate locations to form a mesh-like structure all around the head **(Figure 1A.5**). After the geometry was created, it was rendered against the scalp, to ensure there were no submerged elements. This served as our final quality control check before proceeding to 3D printing.

All headgear prototypes were printed using Form 2 SLA printer (Formlabs, MA, USA) at a layer thickness of 100 microns (**Figure 1B**). The material used was Tough 2000. We ensured that final processing was done in the worldspace of the original scan.

Version 1 included nasion and inion markers along with an offset to compensate for the hair volume expected under a fabric cap (**Figure 1B.2**). Version 2 included an opening at the Cz location to help align/ mark the identified location (with a coil pointer) with respect to the neuronavigation-determined Cz location. This was tested to determine potential sources of headgear error at other scalp locations (i.e. removed from the lDLPFC area) and thereby gauge adjustments needed for the subsequent headgear iteration. This version also included a participant ID marker to prevent mix-up of headgear across test participants. Version 3 incorporated tragus markers to further help position the headgear in accordance with additional anatomical landmarks. Version 4 incorporated additional 10-10 location landmark guides (F3, C3, F4, C4) to further help align the headgear in line with the neuronavigation-guided locations to screen for potential shifts, translations, or scaling errors. The neuronavigation system required positioning two head sensors over the back of the ears via a headband for head movement compensation and to ensure maximum possible measurement accuracy. To prevent interference of the head band with the headgear, dedicated cavities (over each ear) were provided to hold the head tracking sensors in Version 5. These cavities will not be needed for real-world clinical use but were required here to perform measurements and iterate design. The final production design (Version 6) incorporated a material change to balance comfort, flexibility, and robustness (**Figure 5**). For additional functional elements, refer to **Figure 1B**. The final version was fabricated using Selective Laser Sintering (SLS) 3D printing, with a print time of 10.35 hours. A total of 2.82 kg of thermoplastic polyurethane powder was used, including 0.114 kg sintered into the final part. With material priced at $133/kg, the total material cost was $74.90, comprising $14.01 for the final part and $60.89 for excess powder. This process enabled a durable and accurate production of headgear.

### Dummy TMS coil

To facilitate accurate marking of the scalp location directly *underneath* the TMS coil, we 3D printed a figure-of-eight coil with a circular hole coinciding with the isocenter (i.e. the location responsible for initiating the maximum magnetic field). The opening was precisely chosen to allow the experimenter to pass a hand-held pointer / marker through the coil to mark the underlying skull cap. It is standard practice to hold the isocenter right over the brain area that one wishes to stimulate. Further, using a dummy coil, allowed us to account for the coil angle for typical administration.

### Headgear accuracy

Individual MRI data was apriori loaded into Neural Navigator (Brain Science Tools, Utrecht, Netherlands) for neuronavigation-based measurements. The target voxel was set to the individual p46 as determined by the HCP parcellation scheme. The subject was asked to don a close-fitting fabric cap upon arrival, as is standard in clinical settings, to ensure the hair was held as close to the scalp as possible **(Figure 2A**). The individual headgear was then positioned over the cap and the operator confirmed fit. While the headgear by design, conforms to the subject’s head shape and size, and therefore allows minimal adjustment, it is still imperative to cross check that all the anatomical landmarks align with the individual’s anatomy. The p46 scalp location was then marked on the cap using a marker through the dedicated circular opening on the headgear **(Figure 2B**). This location is referred to as ‘hg-p46-s’. The headgear was then removed and a typical setup sequence for a neuronavigation session was followed. Namely, the subjects’ anatomical landmarks were captured via 6 markers (to ensure maximum targeting accuracy) and alignment performed. Upon successful alignment, neuronavigation was initiated. Head movement compensation was always enabled to ensure head tracking and accurate neuronavigation. The dummy TMS coil was then used to navigate to the scalp surface corresponding to the p46 scalp target **(Figure 2D**). A TMS pulse delivery is simulated while holding the dummy coil over the landmark made prior (hg-p46-s), to identify corresponding underlying brain location (hg-p46) **(Figure 2E**). Next, the experimenter navigated to the p46 brain target as identified by the HCP parcellation. A hand-held pointer was positioned through the isocenter hole of the dummy coil to ensure that the navigation beam descending from the pointer intersected the brain target sphere corresponding to p46. The hand-held pointer was held perpendicular to the surface of the head ensuring accurate capturing. The pointer was subsequently removed and the corresponding scalp location (through the isocenter opening) was marked using a different color marker (p46-s). The two marked scalp locations are shown in **Figure 2G** (green: hg-p46-s and purple: p46-s). The two brain target coordinates (blue spheres) are shown in the neuronavigation software in **Figure 2H** (hg-p46 and p46).

**Fig. 2.**
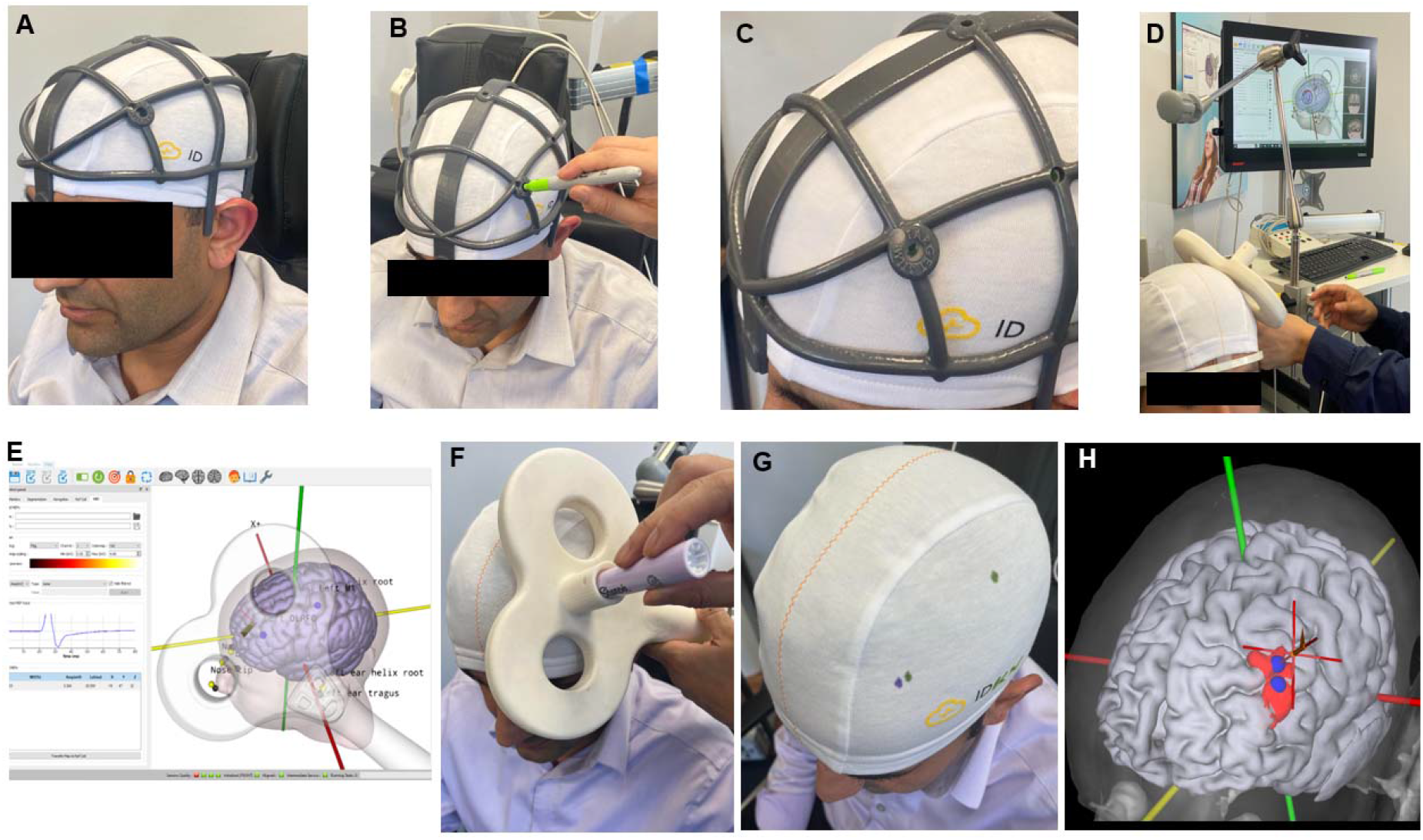
Illustration of testing methodology using the individualized headgear. **[A]** Testing was initiated with the subject donning a classic TMS fabric cap and the subject’s individualized headgear positioned over it. The experimenter ensured proper fit of both the cap and the headgear. While the headgear is designed for proper fit, the experimenter nonetheless confirmed that anatomical landmarks (nasion, tragus, etc.) landed as expected, on the subject. **[B]** The headgear guided scalp coordinate corresponding to p46 (hg-p46-s) was marked on the fabric cap. The experimenter is shown here marking the location with a green pen. **[C]** Close-up of the marked location. The green mark can be seen through the opening on the headgear. **[D]** The headgear is now removed. The subject is now prepped for neuronavigation using subject-specific MRI and dummy coil. **[E]** A TMS pulse delivery is simulated while holding the dummy coil over the green mark (hg-p46-s) to identify corresponding underlying brain location (hg-p46). **[F]** Experimenter navigates to p46 and marks the corresponding scalp location (p46-s) using a purple pen. **[G]** The two marked scalp coordinates are shown (purple:p46-s; green:hg-p46-s). **[H]** The two target brain coordinates are shown in the neuronavigation software (p46 and hg-p46) along with parcel (red). Note the headgear depicted here is version 5 (i.e. one version prior to the final production version). The subject photographed is a study co-author who consented to publication.

All measurements were performed by a trained TMS research scientist (experience:11 years). While numerous measurements were taken as the headgear design was iteratively updated **(Figure 1B)**, a total of 48 measurements were taken on the final version (i.e. 3 targets x 16 subjects).

### Comparison of targets

The euclidean distance between the headgear-navigated p46 and the neuronavigated p46 target was computed and plotted in MATLAB based on each individual’s measured worldspace coordinates. This analysis was repeated for both brain and scalp targets.

### Headgear reproducibility

To test accuracy of repeated application, we mimicked envisioned flow in a clinical setting. The testing was performed by the same operator from the headgear accuracy measurements. On a subset of subjects available to perform such testing (n=8) and using the final production version of the headgear, the operator first positioned a TMS fabric cap using standard procedure. The individualized headgear for the test participant was loaded, location p46 marked on the cap, and the headgear removed. With the fabric cap on, the headgear was repositioned again, and location p46 marked. This sequence was repeated three times in total in the same testing session. To strictly quantify the reproducibility error of the personalized headgear, the fabric cap was intentionally left in place across repeated measurements. The three markings on the cap were made using three different colors **(Figure 7)**. A digital caliper was used to measure the distances between each set of two points. Similar to methodology applied in Mansouri et al., 2018, we determined a centroid of the triangle formed by the set of three points. The mean distance from the centroid was determined.

## Results

Figure 3. depicts individual parcel 46 variability using a subset of subjects highlighting unique and “serpentine” like personalized parcels. The white sphere depicts the identified center of the parcel and was considered the target voxel coordinate for our testing. Note that since the center of the parcel was located in a sulcus for the subject shown in **Figure 3B**, the target voxel was instead placed at the most superficial location possible, as close as possible to the parcel center. **Figure 3** helps reiterate how surface-based reconstruction maintains cortical folds and sulcal banks, which is not possible in volume reconstruction based approaches.

**Fig. 3.**
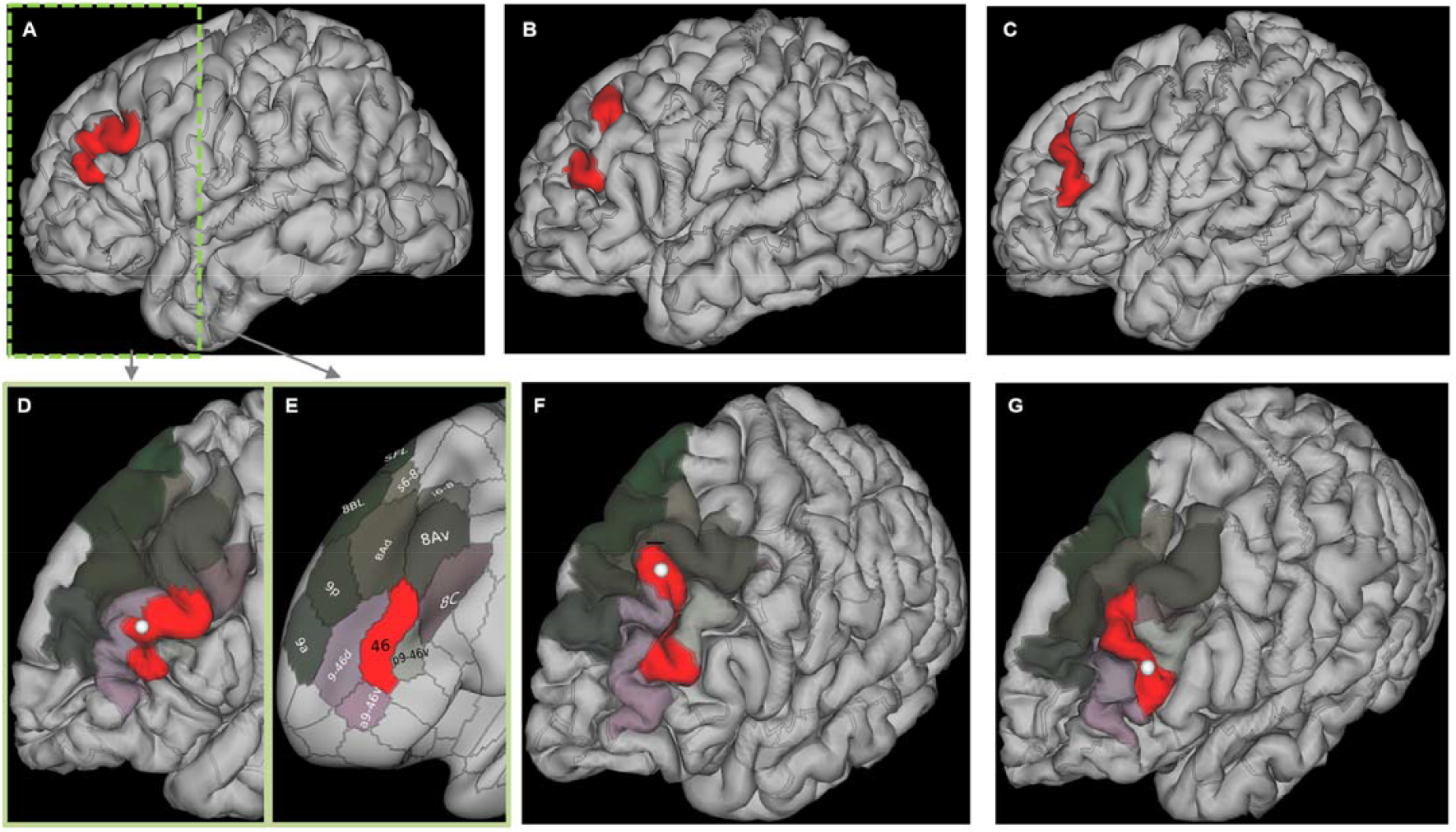
Illustration of individual p46 variability. First row. Individual parcels (red) indicated in individual structural MRI. Data from a subset of subjects from the study (n=3) is shown to highlight variability. **Second row**. The full DLPFC parcellation into 13 areas is shown for the corresponding subjects from the first row. To help identify the 13 regions, an inset from **A** is shown via a corresponding inflated image in **E**. The white sphere (i.e. center of the parcel) indicates the corresponding chosen brain voxel for the testing in this study. Note that since the center of parcel was in the sulci for the subject in **B**, target voxel was set to the most superficial location, close to the center.

While volumetric normalization to MNI space is common in neuronavigated TMS, volumetric structural approaches are based on the original Brodmann brain map that was first published in the early 1900’s and subsequently adapted to MRI using Talairach coordinates mapped onto a population-average MNI brain. Due to inherent variation in sulcal/gyral patterns across individuals, significant smoothing of the cortex is applied in volumetric approaches, leading to loss of targeting precision (Coalson 2018).

Figure 4. depicts comparison of the different brain-to-scalp projection options (Nearest, Normal, and Radial) to the navigated p46 (scalp intersection) location. Data for all three possible approaches was collected in the early phase of the project to identify the most accurate method. Our results indicated that the nearest approach produced the lowest error, and therefore was selected as the projection method of choice for subsequent headgear development.

**Figure 4.**
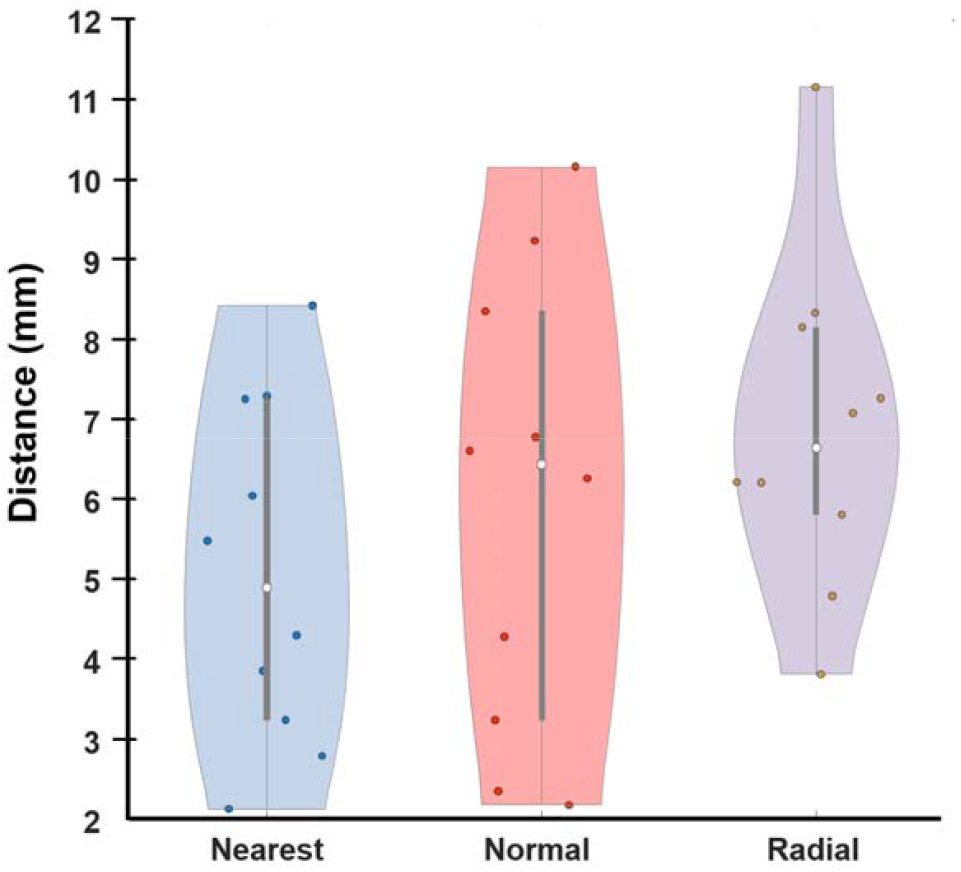
Comparison across different brain-scalp projection methods. Median distances are as follows-Nearest:4.89 mm (IQR:3.24-7.26), Normal:6.44 mm (IQR:3.24-8.35), and Radial: 6.65 mm (IQR:5.81-8.15).

Figure 5. depicts the final embodiment of the individualized headgear for marking p46 on a fabric cap. This production version reflected the culmination of six major iterative design changes to reduce error to the reference method (i.e. the neuronavigation guided coil placement approach) to the extent possible. The final solution is characterized by anatomical landmarks (nasion, left tragus, and right tragus) to guide placement over the skull cap. A circular opening is provided to mark the individual p46 directly on the cap. The final material of choice was Thermoplastic Polyurethane with tensile strength 15 to 20 MPa and was chosen to provide an optimal combination of rigidity and flexibility.

**Figure 5.**
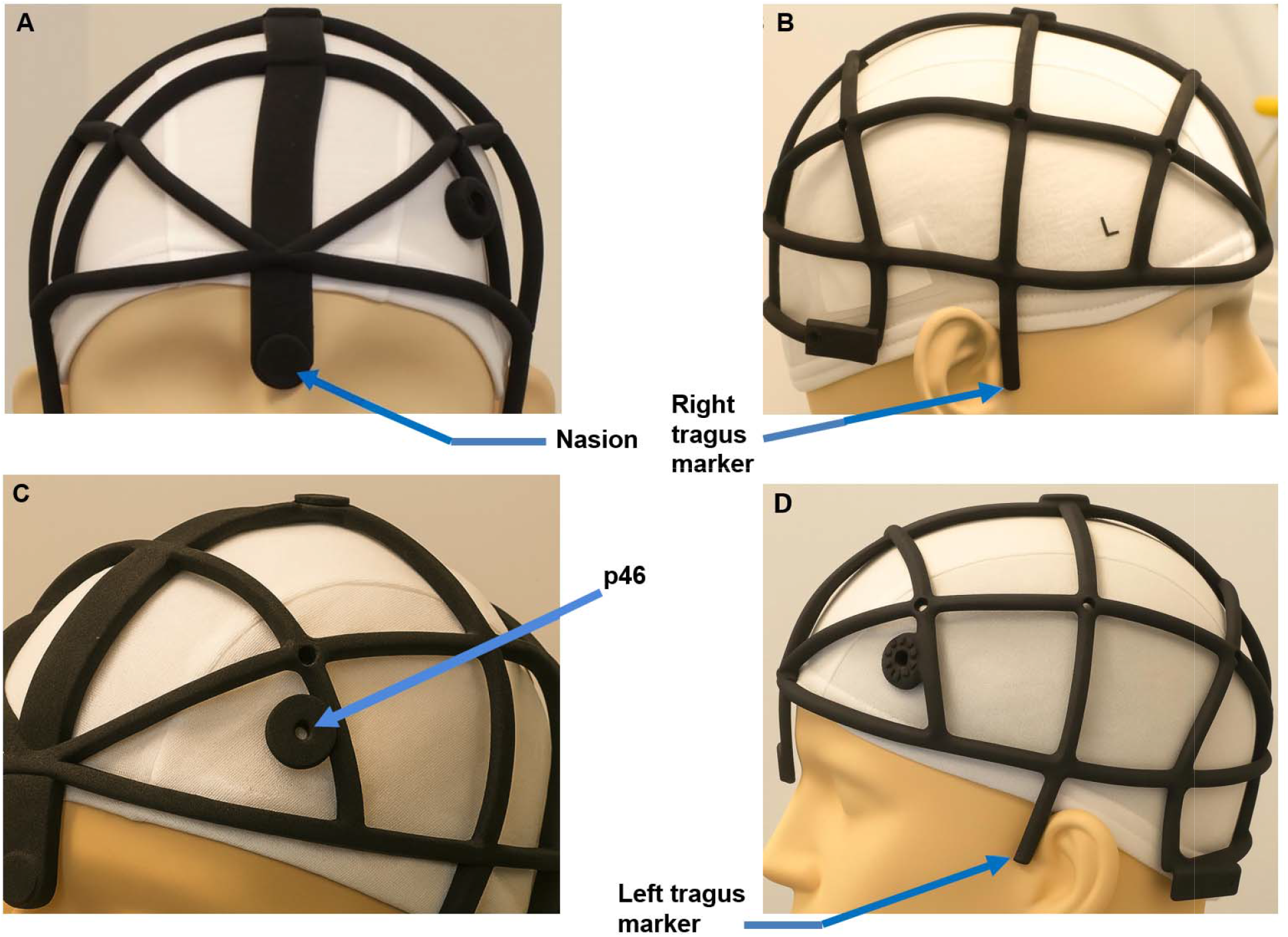
Final embodiment of the individualized p46 headgear. Note that in some sections, the spline segments between adjacent 10-20 positions may not maintain full contact with the scalp .

Figure 6. indicates the variability of the individualized headgear from the neuronavigation-determined location for n=16 subjects. Accordingly, data were unavailable for subjects who could not participate in testing with the final production version of the headgear (i.e. version 6). We note a mean euclidean distance between the individualized and the neuronavigation-guided approach on the brain to be 7.94 mm (SD: 4.99). The corresponding mean euclidean distance between the two approaches on the scalp are noted to be 8.31 mm (SD:4.93 mm).

**Fig. 6.**
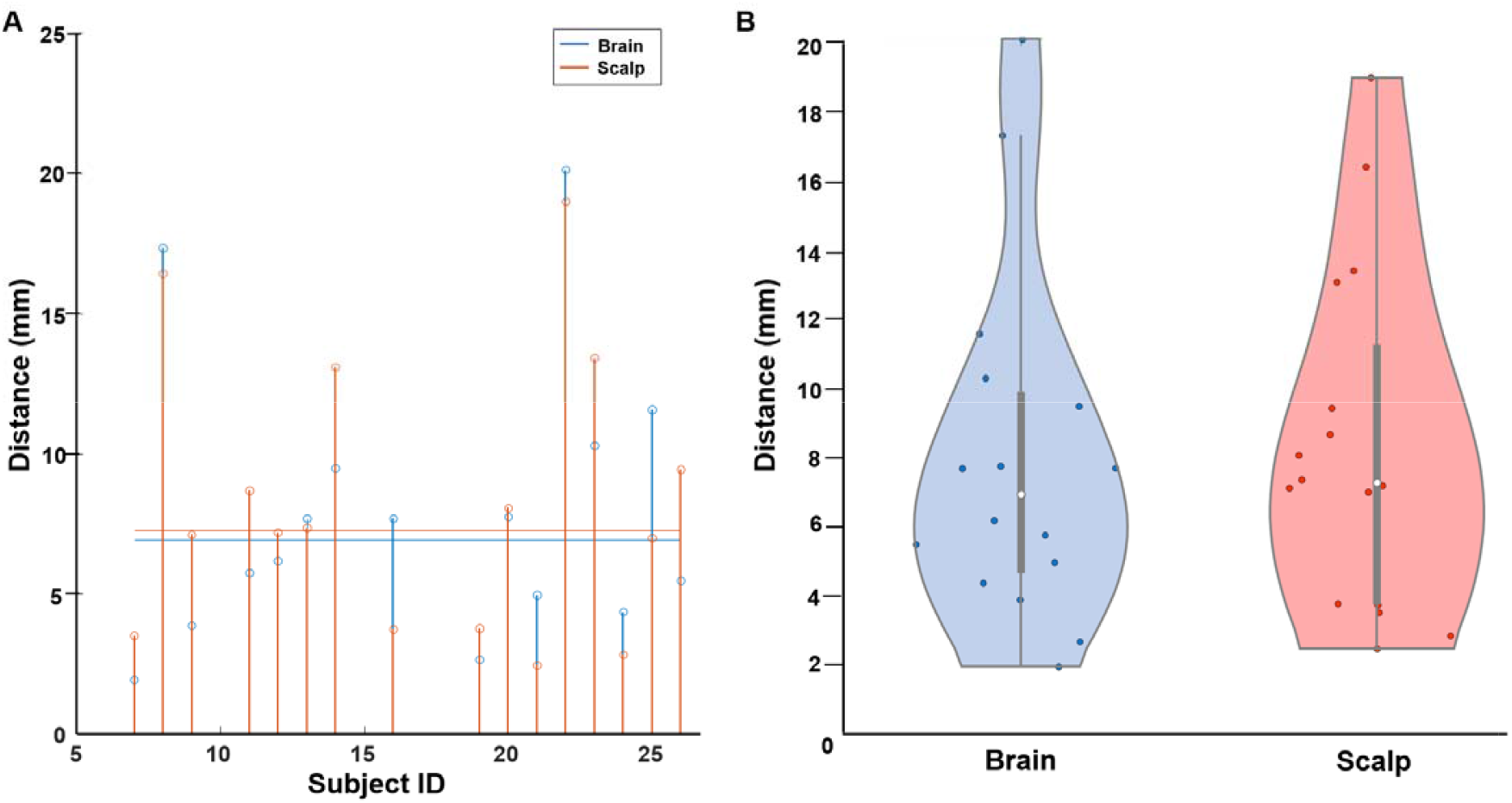
Comparison of individualized headgear determined p46 versus navigated p46. **A**. Distances shown on an individual basis. **B**. Group data (violin) one for scalp and one for brain. Median distance on brain and scalp are 6.93 mm (IQR: 4.66 - 9.89) and 7.27 mm (IQR: 3.74-11.26) respectively.

Figure 7. indicates the result of reproducibility testing. The mean distance from the centroid point was noted to be 1.21 mm (SD: 0.84) across n=8 participants.

**Fig. 7.**
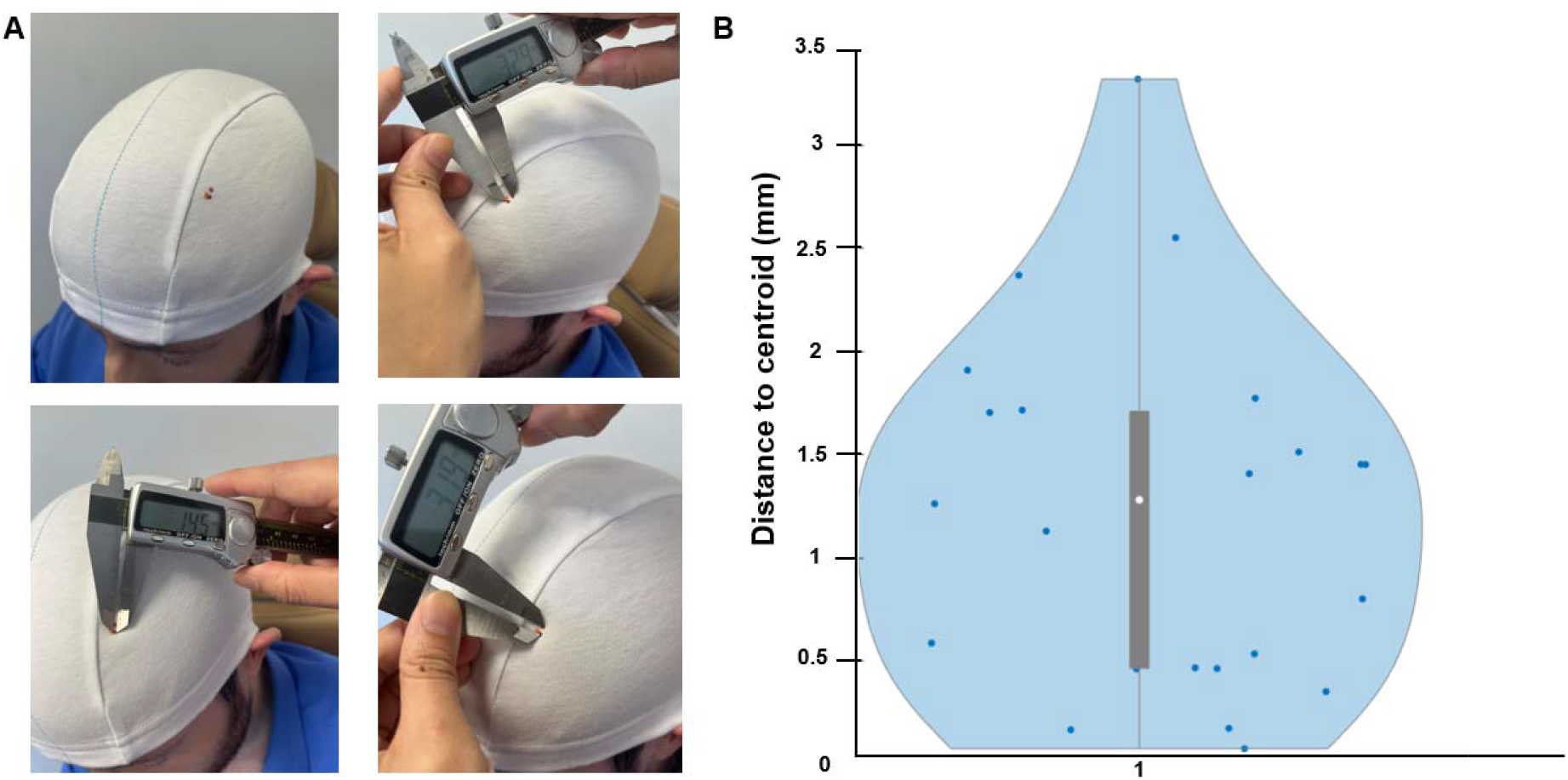
Reproducibility Testing. **A**. Envisioned flow in a TMS center was followed in n=8 subjects. After donning the fabric cap, the individualized headgear for the subject was positioned and p46 scalp location was marked. The headgear was removed and the process was repeated 3 times in total. A caliper was used to measure the distances from any two markings, one at a time. **B**. Test result using a violin plot. Median value noted: 1.28 mm (IQR: 0.46 - 1.71). The subject photographed is a study co-author who consented to publication.

## Discussion

The application of the HCP atlas is beyond the capability of most TMS providers. With the goal of extending HCP-based targeting to community clinical settings, we present an individualized headgear approach that achieves a mean targeting error of ∼ 8 mm relative to the gold-standard navigation method. Given the methodology applied, we note that the error of our solution also includes the error due to neuronavigation-related coil positioning error (conservatively estimated to be ∼3 mm). We therefore estimate the *true* error of our individualized headgear approach to be ∼5 mm and potentially even lower. It is well known that while calibration tolerances for navigation systems are ∼3 mm, additional errors may accrue due to imperfect identification of fiduciary landmarks, co-registration to the MRI, migration of headtracking markers (Mir-Moghtadaei 2022).

We further note inherent potential errors due to scaling and shearing as the navigation system employed in this study uses affine registration instead of rigid body transformation. For instance, hearing protection during MRI scanning, resulted in an un-identifiable helix root in some subjects. Such issues were subsequently relieved by including the medial canthus as a registration landmark and omitting the helix root. However, due to the use of affine registration, errors may have been still introduced that propagated through the neuronavigation process.

The BeamF3 and the 5.5 cm methods are currently considered the preferred coil positioning method for routine clinical use (McClintock 2018). Similar to the BeamF3 approach, we expect practitioners to position a fabric cap as they normally do, and then position the individualized p46 headgear on top of the cap. The practitioner would then proceed to mark the scalp location corresponding to p46 through the circular opening provided on the headgear. The headgear is then removed and the TMS coil is positioned guided by the marking on the cap. The headgear will therefore be used on the first day of the TMS course and not needed again, with the marking on the cap guiding coil placement in subsequent sessions. While BeamF3 is fast, our approach is even faster and easier to use as no measurements and calculations need to be performed. Similar to the BeamF3 approach, the accuracy of repeated targeting is dependent on the practitioner positioning the fabric cap in a repeatable fashion on a day-to-day basis. Our approach is however more expensive but given the goal to make individualized p46 targeting more accessible, is rational. Moreover, any manual scalp-based heuristic is more dependent on operator skill, affecting reproducibility across operators and repeated measurements.

The I-Helmet approach is a relevant virtual neuronavigation approach but requires CT images of the subject’s head, laser scan of the stimulation coil, and practitioner having knowledge of the brain target. While our approach does not provide any guidance with respect to coil orientation, the benefit of our approach is simplicity and easy integration into clinical TMS practices. The off-site HCP processing by our team (to identify brain target) is justified and plans are in place to disseminate 3-D printed individualized headgears in a seamless and rapid fashion. Moreover the core objective of the I-helmet is to provide guidance to the TMS administrator throughout the session, while our headgear is used to identify the scalp target corresponding to the underlying p46 target. With respect to accuracy, the I-helmet approach notes a 3-5 mm positioning error based on deviation from transformation matrices with no validation reported on human participants. While our accuracy is based on a different metric (deviation from neuronavigation determined location), our validation reflects *real-world* errors accounting for relevant factors (skin hardnesses, hair style, etc.). The personalized TMS helmet approach proposed by Badran et al., shares similar core objectives to the I-helmet approach. In addition, the goals were to develop a solution where trained TMS operators are not available or stimulation involves movement (ambulatory, extreme environments, home) and therefore different to our objectives.

As expected, we report high reproducibility as shown with another 3-D printed headset effort (Mansouri 2018). In fact, given the fully encompassing whole-head form-fitted design as opposed to a design focused on the frontal regions (Mansouri 2018), we demonstrate higher reproducibility, as envisioned. Our study is limited by the use of one operator to perform all measurements. Future efforts should test accuracy and reproducibility across multiple operators.

We note future developments of our headgear design. As the focus was to solely help identify the scalp target corresponding to p46, future iterations may explore adding a coil bracket similar to the approach used in Mansouri et al., 2018 to also guide coil orientation. This will however require maintaining the headgear on the fabric cap throughout the treatment and thereby, necessitate making the headgear thinner (with lower profile) and testing for subject comfort over an extended duration application. Further, guidance for the hand area of the motor cortex for each individual could be added. This landmark would serve as the starting point for the sequence of motor threshold determination. The headgear could potentially also take into account induced electric field for every subject (Gomez 2021) and adjust the identified brain target accordingly. These design iterations would be ultimately pursued guided by real-world feedback from clinical centers. As one can expect, these initiatives will be motivated by continued demonstration of the clinical utility (efficacy) of the p46 targeting approach. Finally, our methodology can be translated to any personalized approach including the SAINT approach or analogous targeting of another brain site (for other conditions than Major Depression) when the goal is to make personalized targeting available in community settings.

## Data Availability

All data produced in the present work are contained in the manuscript

## Acknowledgements

This research was supported by funding from NIH NIMH R44MH126833.

